# Characterizing Preschool Children’s Multi-Matrix Air Pollutant Exposures Across Home and Early Childhood Education Settings: A Paired Silicone Wristband Study Protocol

**DOI:** 10.64898/2026.08.27.26361470

**Authors:** Abby D. Mutic, Linda A. McCauley, Angelina Andrew, Anne M. Fitzpatrick

**Affiliations:** Nell Hodgson Woodruff School of Nursing, Emory University, Atlanta, GA; University of North Carolina at Chapel Hill, Chapel Hill, NC; Department of Pediatrics, Emory University School of Medicine, Atlanta, GA

**Keywords:** Exposure assessment, Micro-environment, Indoor Air Pollution, Pediatrics, Asthma, Learning Environment

## Abstract

**Background:** Children spend more than 90% of their time indoors, and early childhood education settings (ECEs) are an understudied, high-occupant-density indoor microenvironment where exposure to volatile organic compounds, particulate matter, and other toxicants has been documented. Limited knowledge exists on ECE-specific exposures affecting young children and how they compare to exposures in the home.

**Methods:** This prospective, repeated-measures pilot study targeted enrollment of 44 preschool-aged children and 8 ECE staff across two geographically and sociodemographically distinct ECEs in metropolitan Atlanta, Georgia. Paired silicone wristbands, one home-designated and one ECE-designated, were exchanged between settings across three consecutive days and nights beginning at enrollment to characterize microenvironment-specific exposure. A single spot urine sample was also collected from each child. Continuous indoor air quality monitoring was conducted in two classrooms per site. Caregivers and ECE staff completed structured questionnaires assessing home and ECE environmental characteristics, child respiratory risk, and protocol feasibility and acceptability. Feasibility was evaluated using eight pre-specified indicators spanning recruitment and enrollment, wristband wear duration and loss by microenvironment, urine sample collection completeness, and survey completion by instrument and respondent group.

**Conclusion:** This pilot will establish feasibility and acceptability parameters for a paired, multi-matrix silicone wristband protocol across home and ECE microenvironments. Findings will inform the design, sample size, and power calculations for a subsequent study testing indoor air interventions and pediatric respiratory outcomes in ECEs. Feasibility outcomes are reported in a companion manuscript.

## 1. Introduction

Children spend more than 90% of their time indoors, and their physiology places them at disproportionate risk from the pollutants they encounter there. Children breathe more rapidly, inhale a greater volume of air relative to body size, and have respiratory, neurological, endocrine, and immune systems that are still developing [1,2]. Indoor pollutant concentrations frequently exceed those measured outdoors. Additional circumstances compound this risk for some children including household tobacco smoke exposure, residence in low-income or inner-city neighborhoods, and high traffic locations [3-5]. Proximity to industrial or chemical facilities adds further risk, especially in areas where communities of color have disproportionately experienced discriminatory zoning, redlining, and land-use practices [5]. Respiratory illness and disease symptoms such as difficulty breathing, cough, and wheezing also disproportionately affect ethnic minority children with Black children, specifically, having a higher prevalence of asthma, asthma-related hospitalizations, and a higher risk of death compared to other racial groups [6].

Early Childhood Education Settings (ECEs) are a particularly understudied indoor microenvironment. Toxicants including brominated flame retardants, bisphenol-A, phthalates, volatile organic compounds (VOCs), particulate matter, and pesticides have been identified in childcare settings, frequently at concentrations that raise safety concerns [7–15].Further, ECE facilities routinely house up to four times the occupant density of comparably sized office buildings [16], yet little regulatory attention has been paid to the disparate exposure risks this creates across urban and rural settings [17, 18].

### 1.1 Health Disparities and Exposures in Childcare Settings

This exposure burden compounds existing health disparities. Studies of ECEs in California have documented elevated VOC concentrations, including chloroform, benzene, and formaldehyde, a known human carcinogen [10,19]; comparable inner-city childcare facilities in Washington, DC have reported VOC concentrations more than 14 times higher than those measured in Northern California facilities [11], suggesting that geographic and community context shape ECE exposure profiles as much as facility-level characteristics do. Despite this, the majority of pediatric exposure research has focused on prenatal or home environments, leaving the ECE setting where many children spend the greater part of their waking hours, comparatively unexamined. This gap is particularly consequential because home and ECE microenvironments differ in ways that are likely to shift a child’s total exposure burden. For example, ECE facilities contain more textiles, carpet, and shared items than most homes. Under-resourced centers may be less able to perform routine maintenance, repair structural damage, or manage moisture and pest issues that contribute to indoor contamination. At the same time, high-quality early care and education can independently support children’s health and development: center-based care and higher-quality childcare, including greater caregiver sensitivity, stimulation, and structured programming, have been associated with better developmental outcomes even after accounting for socioeconomic and home-environment factors [20], underscoring that ECE quality is not only a source of exposure risk but also a modifiable protective factor worth characterizing alongside it. No existing study has directly compared the same child’s exposure across both the home and ECE microenvironments, and research is lacking on ECE-specific exposures in the Southeastern United States.

### 1.2 Passive Exposure Assessment Via Silicone Wristbands

Current exposure assessment methods are poorly suited to closing this gap in ECE and home settings alike. Biomarker measurement in urine, blood, or breast milk requires biological sample collection while active air-sampling devices require a pump, battery, and collection apparatus [21]. Both methods are impractical to implement with young children and many of the devices are cumbersome and noisy for use in school settings. Silicone wristbands offer a passive alternative. They are low-cost, noninvasive, and easy to use, and have demonstrated detection frequencies comparable to active sampling devices, with better correlation to urinary biomarkers in at least one validation study [22,23]. Wristbands have been deployed successfully across a range of ages and settings, including with preschool-aged children [24– 27], but their use specifically within childcare settings remains emergent, and no study has used a paired wristband design to characterize the same child’s exposure across home and ECE settings concurrently.

### 1.3 Specific Aims

This protocol describes a pilot study designed to address the gap in the literature regarding the utilization of wristbands in ECE settings. The primary aim is to assess the feasibility of a paired silicone wristband protocol as a method for characterizing preschool children’s air pollutant exposures within a microenvironment. Specifically, this study addresses: (RQ1) To what extent is the wristband protocol acceptable to enrolled children and ECE staff; and (RQ2) to what extent is the protocol practical to implement across home and ECE settings, including the rate of successful sample and data collection.

Findings from this pilot will directly inform a study assessing indoor air exposures, filtration interventions, and pediatric respiratory outcomes.

## 2. Methods

### 2.1 Study Design

This was a prospective, repeated-measure, observational pilot study without a randomized or comparison arm approved through Emory IRB. Each enrolled child and ECE staff member served as their own comparison across the home and ECE microenvironments over three consecutive study days (Fig. 1). This protocol is described as multi-matrix because it pairs data from multiple sample types, including silicone wristbands, urine, and indoor air monitoring, and multi-microenvironment because these samples were collected in parallel across both home and ECE settings.

**Figure 1.**
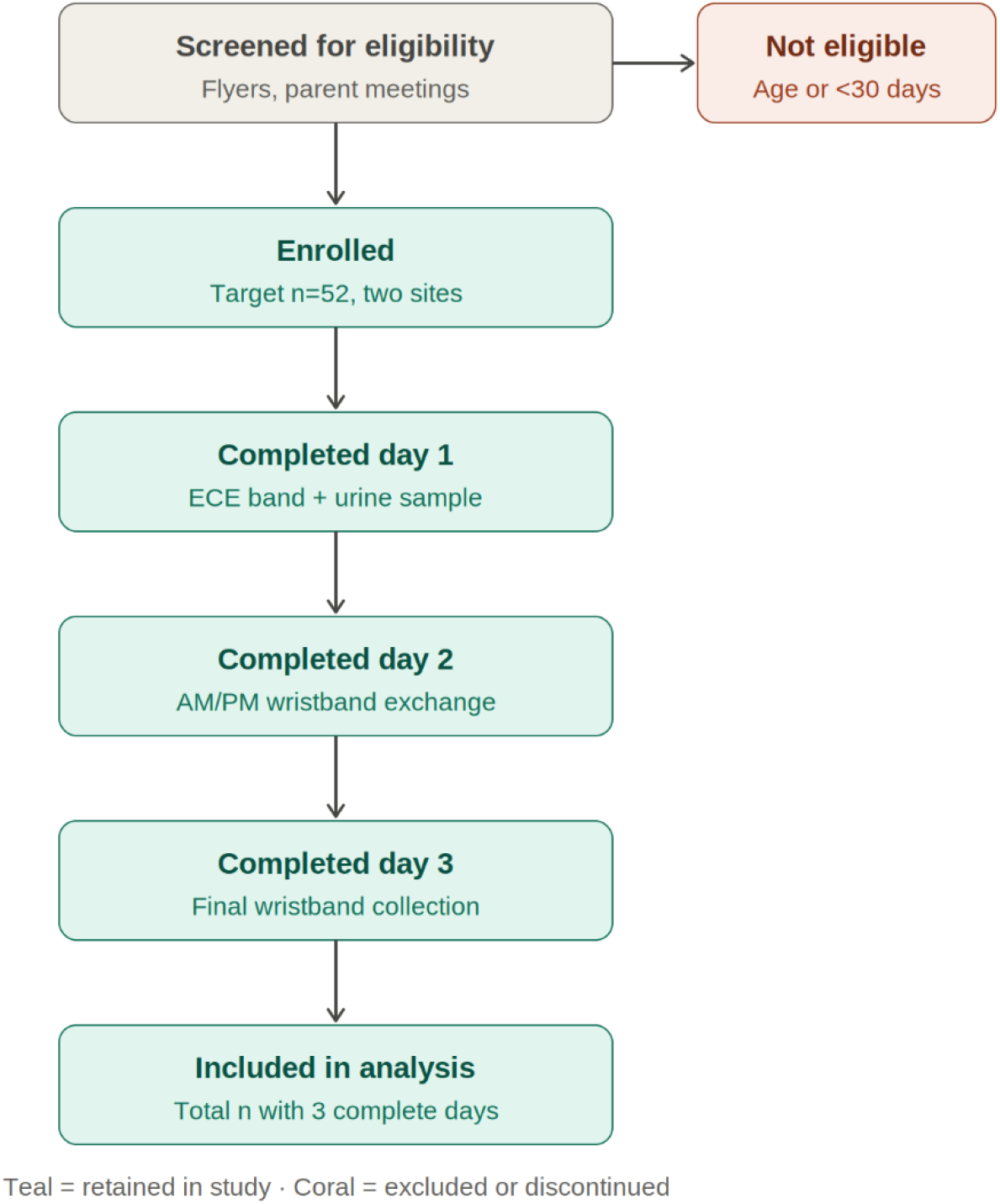
Participant flow diagram, enrollment through completion. Initial image generated using Anthropic / Claude, Sonnet 5 and subsequently edited by the authors. All elements were reviewed for accuracy and completeness.

### 2.2 Setting and Community Partnership

This study was conducted in partnership with Sheltering Arms, Georgia’s largest nonprofit early childhood education organization, which provides early education, care, and family support services to more than 3,600 children and families across 16 metropolitan Atlanta ECEs. Sheltering Arms serves a demographically diverse population, the majority of whom are Black (77%) and living at or below the federal poverty line (50%). Our research team has provided health screenings, physical examinations, and health education to Sheltering Arms families for many years and this study leverages that established relationship to conduct community-engaged participatory research. For this study, Sheltering Arms facilitated initial communication with each site’s school administration and helped arrange an informational parent meeting for interested families at each center.

Data collection occurred across two Sheltering Arms centers selected to represent contrasting community and environmental contexts to compare urban and suburban characteristics. These two catchment areas differ meaningfully across dimensions with established relevance to pediatric environmental exposure.

Urban ECE zip code is characterized by an older housing stock (median construction year 1958, with roughly 37% of homes built before 1950), higher median household income (∼$108,000), and a smaller, majority-White, majority-owner-occupied population (2 persons/household on average; 68% owner-occupied). Suburban ECE catchment area, by contrast, reflects a substantially newer housing stock (median construction year 2001, with fewer than 1% of homes built before 1950), a majority-Black population, a lower median household income (∼$79,000–95,000 depending on geographic level), larger average household size (∼3.3 persons/household), and a higher proportion of renter-occupied housing (52%). Because older housing stock carries greater potential for lead paint and other structural-material exposures, while population density and proximity to urban traffic corridors shape ambient air pollutant exposure, this purposive two-site design was intended to capture meaningful community-level heterogeneity that may modify ECE-based exposure risk, complementing the individual-level, within-child home-ECE comparison at the core of the wristband protocol. Each center’s cohort completed a separate three-day study period. All participant-facing procedures occurred either at the ECE or in participants’ homes. Collected biological and environmental samples were transported to and processed at the Emory University LEADER laboratory.

### 2.3 Recruitment and Participant Eligibility

Children and ECE staff were recruited via convenience sampling at both participating facilities using study flyers and informational materials posted in ECE lobbies, take home materials distributed to classroom families, through teacher-assisted parent meetings, and by having a study table set up in the ECE lobby. Classrooms were selected by the ECE administrator as they represented the only classrooms serving preschool-aged (3–5 year old) children at each site. ECE staff assisted the research team by connecting families with study personnel through email listservs, in-person discussion, and distribution of study materials. Interested participant families were directed to meet the study team in the morning during child dropoff times or after school during pickup times and were screened using an eligibility checklist. ECE staff and teachers were recruited directly from the participating classrooms, and eligibility was determined by the study staff.

Study inclusion criteria included 1) children aged 3–5 years enrolled in one of the two participating Sheltering Arms ECEs for at least 30 days prior to enrollment, duration selected to accommodate the half-lives of most target toxicants, and 2) ECE staff who had been employed at the facility for at least 30 days and who spent the majority of their workday indoors at the study site. Children with a documented chronic liver or kidney condition and children with a primary language other than English or Spanish were excluded. The study targeted enrollment of 52 participants (44 children and 8 ECE staff) across the two sites, anticipating up to 10% attrition or incomplete data. All recruitment and study materials were available in English and Spanish, and bilingual research assistants were available as needed; translated materials were back translated to English for quality control.

### 2.4 Multi-Microenvironment Wristband Protocol

Each enrolled child and staff participant wore paired silicone wristbands, exchanged between the ECE and home settings, over the study period of three consecutive nights and school days. At enrollment, a study team member (a CITI-trained research assistant or the study principal investigator) placed a home color-designated wristband on the child, worn overnight until arrival at the ECE the following morning. At arrival on day 1, the home wristband was removed and exchanged for an ECE color-designated wristband, worn without removal throughout the school day. At pickup, the ECE wristband was exchanged for the home wristband, worn overnight until the following morning. This morning and afternoon exchange was repeated with ECE staff or study team assistance on days 2 and 3, so that the home wristband was worn for three consecutive nights in total. On the morning of day 3, the home wristband was exchanged for the ECE wristband for a final day of ECE wear. At pickup on day 3, the ECE wristband was collected and stored, ending wristband wear for the study period. When not in use, each removed wristband was placed immediately into an individually labeled, sealed storage bag identified by subject ID and held on site by the study staff. All wristbands were transferred to the Emory LEADER laboratory at the end of the study period, where they were processed and stored at −20°C pending extraction and analysis (Fig. 2).

**Figure 2.**
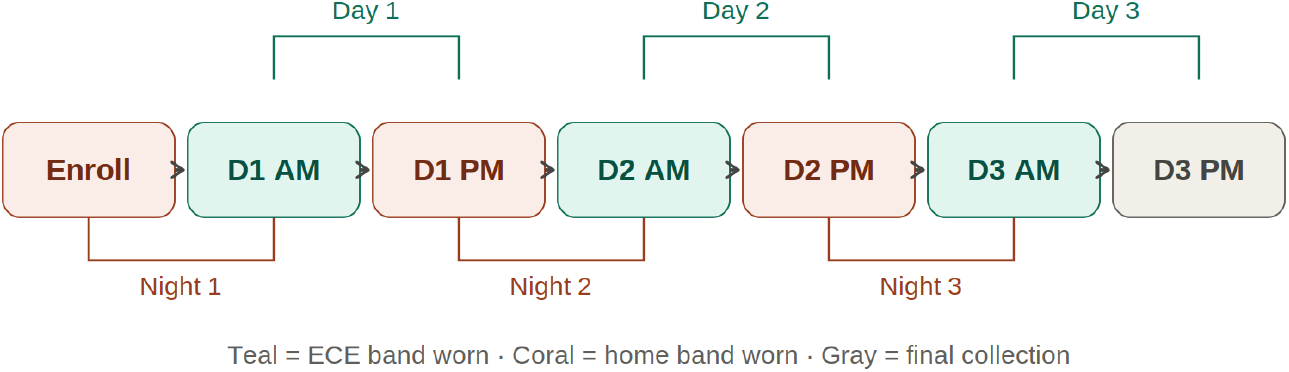
3-day AM/PM wristband exchange. Draft created with Anthropic / Claude, Sonnet 5. The final figure was refined and verified by the authors.

### 2.5 Biological Sample Collection

A single urine sample (10–30 mL) was collected on day 1 of the study period from each child participant via self-collection (with caregiver assistance as needed) into a provided specimen collector and transferred to a sterile biospecimen cup labeled with subject ID. Participants and caregivers were given the option of having a parent/guardian, ECE staff member, or research team member assist with collection, according to family preference. Samples were stored locally on ice, transported to the Emory LEADER laboratory at the days end, aliquoted in the laboratory, and stored at −20°C pending extraction and analysis.

### 2.6 Environmental (Classroom) Air Quality Monitoring

Continuous indoor air quality monitoring was conducted in the two classrooms at each participating center previously identified by the ECE administrator and where the enrolled children were attending (four classrooms total: two at suburban ECE and two at urban ECE). Indoor air quality was measured in each classroom using individual AirThings View Plus monitors (Airthings AS, Oslo, Norway), a seven-sensor device that measures radon, particulate matter, total volatile organic compounds (VOCs), humidity, temperature, noise, and carbon dioxide (CO2) and converts these readings into actionable indoor air quality metrics [28]. Monitors were deployed for a minimum of 14 consecutive days per classroom, a duration selected to allow the comparison of air quality during periods when children were present during school hours against periods when the building was unoccupied (after-school hours, overnight, and weekends). At each ECE site, one classroom was equipped with an AirThings View Plus sensor paired with a hub, and a second classroom was equipped with a sensor only, which connected wirelessly to the paired hub. Devices were placed on desks or shelving at waist-to-shoulder, the manufacturer’s recommended “breathing zone”, out of children’s reach and sight, and away from doors, windows, vents, and heat sources. Hubs were installed first and connected to the cloud before sensor installation, allowing sensors to automatically connect to the nearest hub once powered on [29]. Air quality data and real-time notifications provided a room-by-room dashboard with live readings and long-term trend graphs that allowed direct comparison across classrooms at the same site.

### 2.7 Survey Instruments

Four survey instruments were administered across the study period (Table 1). The Home Environment Questionnaire was completed by the child caregiver and returned within the 3-day study period. Data ascertained personal care, cleaning, and pesticide product use in and around the home; recent construction, remodeling, or structural damage; floor type; and heating, cooling, and ventilation practices, alongside family demographics (household size, income, poverty/income ratio, race/ethnicity, education, marital and insurance status) and child health history [15].

**Table 1.** Study Survey Instruments.

| **Instrument** | **Domain / content** | **Respondent** | **Timing** |
| --- | --- | --- | --- |
| **Home Environment Questionnaire** | Personal care, cleaning, and pesticide product use; recent construction, remodeling, or structural damage; floor type; heating, cooling, and ventilation practices; household demographics (size, income, poverty/income ratio, race/ethnicity, education, marital and insurance status); child health history | Parent/guardian<br>N=44 | Day 1 (completed on-site or sent home and returned by day 3) |
| **ECE Facility Questionnaire** | Personal care, cleaning, and pesticide product use; construction, remodeling, and structural condition; heating, cooling, air filtration, and ventilation practices; daily child/staff occupancy; children's time-use patterns across classroom, bathroom, outdoor, and other building areas | Classroom teacher and school administrator<br>N=8 | During the 3-day study period |
| **Health Questionnaire (CHART)** | Respiratory symptoms and asthma risk (wheeze and cough episodes, medication use, healthcare utilization in prior 12 months); categorized as low, moderate, or high risk | Parent/guardian<br>N=44 | Once, during the 3-day study period |
| **Feasibility Survey** | Perceived ease and acceptability of the wristband exchange procedure, urine collection, and questionnaire burden; broader participant concerns about environmental exposures and children's health and development | Parent/guardian and ECE staff<br>N=52 | Day 3 (final day of study period) |

*CHART = the respiratory screening tool used to assess asthma risk.*

The ECE Facility Questionnaire ascertained comparable information at the facility level — personal care, cleaning, and pesticide product use; construction, remodeling, and structural condition; and heating, cooling, air filtration, and ventilation practices — along with staff to child ratio and children’s time-use patterns across classroom, bathroom, outdoor, and other building areas [15]. The Health Questionnaire was administered once during the three-day study period using the CHART screening tool [30] to assess respiratory symptoms and asthma risk, categorizing each child as low, moderate, or high risk based on parent- or guardian-reported wheeze and cough episodes, medication use, and healthcare utilization in the prior 12 months. The Feasibility Survey [31] was administered to participating families and ECE staff at the conclusion of the study period (day 3) and captured the perceived ease and acceptability of the wristband exchange procedure, urine collection, and questionnaire burden, along with participants’ broader concerns about environmental exposures and children’s health and development. All four instruments were paper based to allow for caregiver and flexibility and choice to complete the surveys on site or to take them home and return before the study completion. Completed paper instruments were manually entered into REDCap by trained research assistants on a weekly basis throughout the study period and original copies stored in a secured lockbox.

### 2.8 Planned Data Analysis

Wristband and urine samples will be analyzed using untargeted metabolomics to characterize the range of chemical airborne exposures captured across the home and ECE microenvironments. Feasibility will be evaluated using eight pre-specified indicators spanning recruitment and enrollment rates, wristband wear duration and loss by microenvironment, urine sample collection completeness, and survey completion by instrument and respondent group. These indicators are intended to establish preliminary parameters of exposure concentrations and magnitude of difference between home and ECE settings to inform the design and power calculations for a subsequent, larger-scale study. The exploratory data analysis methods allow for hypothesis generating when there is a limited understanding of routine ECE-specific exposures and implications for child health [17].

### 2.9 Study Procedures and Data Management

Study data were stored using the Research Electronic Data Capture (REDCap) system, encrypted for storage on collection devices and transmitted to secure Emory University servers. Data were de-identified using a study ID number wherever possible. Study consents were stored separately from the assigned study IDs. No video recordings or participant names appear in study records shared outside the immediate research team. De-identified data may be placed into public repositories, where researchers must execute a data use agreement prior to access.

### 2.10 Compensation

Participating families received a $50 gift card upon completion of the full study protocol, including the questionnaire battery, wristband exchanges, urine sample collection, and the feasibility survey. Participating ECE classroom teachers received a $100 gift card in recognition of their active role in recruitment, family communications, urine sample collection support, wristband exchange assistance, and completion of the ECE facility and feasibility questionnaires over the study period. Each Sheltering Arms ECE received an EnviroKlenz air filtration unit donated by Timilon Corporation for use in any classroom [32]. This pilot did not evaluate filtration performance or air quality measures in conjunction with the EnviroKlenz unit.

## Discussion

This protocol describes a novel paired-microenvironment approach to characterizing young children’s environmental air and chemical exposures, using passive silicone wristband sampling exchanged between home and ECEs. The procedures collected data from multiple matrices to capture complex environmental exposures and risk behaviors associated with exposure [33]. Air pollutant mixtures will be estimated using wearable wristbands, urine biomarkers, and coupled with indoor air quality measurements. The structured survey instruments administered to caregivers, ECE staff, and facility leadership allows direct within-child comparison of exposure burden and risk across the two microenvironments where preschool-aged children spend the majority of their waking hours. To our knowledge, no prior study has applied this paired design specifically within childcare settings, and none has characterized ECE-specific chemical exposures in the Southeastern United States.

### 2.2. Strengths

Several design features strengthen this protocol’s contribution. First, the community partnered research with Sheltering Arms established through several years of collaborative health programming and guidance on important health-related questions in the community provided infrastructure for the study aims and design, recruitment, family trust, and facility access. Research partnerships with this level of trust and buy-in are difficult to replicate through cold outreach, particularly among historically under-resourced populations [34]. Second, the purposive selection of two centers differing meaningfully in neighborhood income, racial and ethnic composition, housing age, and urban/suburban density allows us to characterize how community-level context may shape ECE-based exposure risk which complements the individual-level and provides contrast to the home environment. Third, reliance on passive, low-burden sampling methods (wristbands and single-void urine collection rather than active air sampling or blood draws) is developmentally appropriate for preschool-aged children and minimizes disruption to normal ECE operations, a frequently cited barrier to research in childcare settings [17]. Fourth, the paired wristband design, in which the same wristband was exchanged between each child’s home and ECE environments, enables direct within-child comparison of exposure across both microenvironments, an approach no prior childcare-based study has applied.

### 2.2. Limitations

This protocol also has anticipated limitations that should inform interpretation of the forthcoming feasibility results. As an unblinded, observational pilot without a comparison arm, it is designed to establish feasibility parameters such as recruitment and retention rates, wristband wear duration, sample and survey completeness rather than to test hypotheses about exposure-outcome relationships. Likewise, the untargeted metabolomics approach helps to formulate research questions, however, it does not permit confirmatory statistical inference at this stage. Convenience sampling at two centers, with a target enrollment of 52 participants, limits generalizability but is important to establish protocol viability and allows us to anticipate needed resources, time, and future constraints [35]. Another limitation is the short study window at each site does not capture seasonal or day-to-day variability in either the home or classroom environments. Given the location of the study, the addition of other environmental factors such as heat, humidity, and pollen allergy counts would better characterize indoor air pollutants and exposure variability even if used as controls. Finally, reliance on caregivers and ECE staff to provide accurate self-report for the Home Environment Questionnaire, ECE Facility Questionnaire, and CHART respiratory screening introduces the possibility of recall or social desirability bias, though this is a well-known limitation in health behavior surveys and is partially mitigated by pairing the self-report with objective biomarker data [36].

## 3. Conclusion

With more emphasis placed on structured preschool environments, it is critical to identify and maintain key features of healthy learning environments to reduce absenteeism and optimize child health and development. Findings from this pilot will directly inform the design, sample size, and power calculations for a subsequent, larger-scale study examining indoor air interventions and pediatric respiratory outcomes in ECEs. By establishing the feasibility and acceptability of a paired wristband protocol across home and ECE microenvironments, multi-matrix environmental and biological samplings methods, and by purposively sampling across differing communities, this pilot lays methodological and community-partnership groundwork for a rigorous explanation of early child environmental exposures.

## Data Availability

All data produced in the present study are available upon reasonable request to the authors.

## Acknowledgements

We would like to thank all the patients and families who have participated in this study. We would also like to thank our collaborators at the Sheltering Arms study sites. This time and willingness to participate in children’s health research make this possible.

## Data availability

No data was used for the research described in the article.

## Author Contributions

### CRediT

**Abby Mutic:** Writing-original draft, review and editing, Conceptualization, Methodology, Funding acquisition. **Linda McCauley**: Writing-review and editing, Conceptualization, Funding acquisition. **Angelina Andrew**: Writing-review and editing. **Anne Fitzpatrick**: Writing-review and editing, Supervision, Methodology.

## Ethics Approval

The study received approval from the Emory University Institutional Review Board (IRB00112664).

## Declaration of generative AI and AI-assisted technologies in the manuscript preparation process

During the preparation of this work the authors used Anthropic / Claude, Sonnet 5 to develop and color code the workflow and wristband exchange figures. After using this tool/service, the authors reviewed and edited the content as needed and take full responsibility for the content of the published article.

